# Diarrhea prevalence in a randomized, controlled prospective trial of point-of-use water filters in homes and schools in the Dominican Republic

**DOI:** 10.1101/2020.10.21.20217299

**Authors:** Nathan Tintle, Kristin Van De Griend, Rachel Ulrich, Randall D. Wade, Tena M. Baar, Emma Boven, Carolyn E. A. Cooper, Olivia Couch, Lauren Eekhoff, Benjamin Fry, Grace K. Goszkowicz, Maya A. Hecksel, Adam Heynen, Jade A. Laughlin, Sydney M. Les, Taylor R. Lombard, B. Daniel Munson, Jonas M. Peterson, Eric Schumann, Daniel J. Settecerri, Jacob E. Spry, Matthew J. Summerfield, Meghana Sunder, Daniel R. Wade, Caden G. Zonnefeld, Sarah A. Brokus, Francesco S. Moen, Adam D. Slater, Jonathan W. Peterson, Michael J. Pikaart, Brent P. Krueger, Aaron A. Best

**Affiliations:** Department of Mathematics and Statistics, Dordt University, 700 7^th^ St. NE, Sioux Center, IA 51250; Department of Sociology, Dordt University, 700 7^th^ St. NE, Sioux Center, IA 51250; Department of Mathematical Sciences, Montana State University, P.O. Box 172400, Bozeman, MT 59717; Biology Department, Hope College, 35 E. 12^th^ St., Holland, MI 49423; Chemistry Department, Hope College, 35 E. 12^th^ St., Holland, MI 49423; Department of Biology, Dordt University, 700 7^th^ St. NE, Sioux Center, IA 51250; Geological and Environmental Sciences Department, Hope College, 35 E. 12^th^ St., Holland, MI 49423

**Author notes:** **Corresponding Author:** Aaron A. Best.

**Keywords:** Drinking Water, Point-of-Use Filter, 16S rRNA Community, Diarrhea, Heavy Metals

## Abstract

**Background:** Lack of sustainable access to clean drinking water continues to be an issue of paramount global importance, leading to millions of preventable deaths annually. Best practices for providing sustainable access to clean drinking water, however, remain unclear. Widespread installation of low-cost, in-home, point of use water filtration systems is a promising strategy.

**Methods:** We conducted a prospective, randomized, controlled trial whereby 16 villages were selected and randomly assigned to one of four treatment arms based on the installation location of Sawyer^®^ PointONE™ filters (filter in both home and school; filter in home only; filter in school only; control group). Water samples and self-reported information on diarrhea were collected at multiple times throughout the study.

**Results:** Self-reported household prevalence of diarrhea decreased from 25.6% to 9.76% from installation to follow-up (at least 7 days, and up to 200 days post-filter installation). These declines were also observed in diarrhea with economic or educational consequences (diarrhea which led to medical treatment and/or missing school or work) with baseline prevalence of 9.64% declining to 1.57%. Decreases in diarrhea prevalence were observed across age groups. There was no evidence of a loss of efficacy of filters up to 200 days post filter installation. Installation of filters in schools was not associated with decreases in diarrhea prevalence in school-aged children or family members. Unfiltered water samples both at schools and homes contained potential waterborne bacterial pathogens, dissolved heavy metals and metals associated with particulates. All dissolved metals were detected at levels below World Health Organization action guidelines.

**Conclusions:** This controlled trial provides strong evidence of the effectiveness of point-of-use, hollow fiber membrane filters at reducing diarrhea from bacterial sources up to 200 days post installation when installed in homes. No statistically significant reduction in diarrhea was found when filters were installed in schools. Further research is needed in order to explore filter efficacy and utilization after 200 days post-installation. Trial registration: ClinicalTrials.gov, NCT03972618. Registered 3 June 2019 - Retrospectively registered, https://clinicaltrials.gov/ct2/show/NCT03972618.

## Background

Globally, diarrhea caused over 1.65 million deaths in 2016 [1] and half a million deaths in 145 low- and middle-income countries in 2012 due specifically to inadequate drinking water [2]. According to the World Health Organization (WHO), there are 1.7 billion cases of childhood diarrheal disease annually and 525,000 children under 5 die from the disease each year, making diarrheal disease the second leading cause of death in children under 5 [3]. Since a major source of diarrhea is fecal pathogens via fecal-oral transmission [4, 5] many of these lives could have been saved through clean drinking water [2] and proper hand hygiene [5, 6].

The Dominican Republic is considered a middle-income country [7], and thus is at potentially high risk for negative impact of diarrheal disease. Results of household surveys confirm high prevalence of diarrhea in children under 5 in the Dominican Republic (32.3% treated for diarrhea with oral rehydration salts in 2002; 46.3% in 2007 and 52.8% in 2013) [8]. Other studies have found a similarly high burden of diarrheal disease. In 2003 the Dominican Republic Demographic and Health Survey reported that an average of 14% of all children under five years suffered from diarrhea, with rates up to 29% in certain provinces [9].

Other studies have found related concerns about sustainable access to clean drinking water. In a peri-urban district of Santo Domingo, a study of 266 households found that although 57% of interviewees believed their child to be at risk for diarrhea, and 90.6% believed that boiling water could prevent diarrhea, only 42% reported that it was rare for their child to drink untreated water with 34.5% stating ‘insufficient fuel’ as the primary barrier [10]. Another study conducted in the Puerto Plata region to evaluate *E. coli* levels in household water sources found that in unimproved water sources, 47% were of high to very high risk according to WHO water quality guidelines and in improved sources, 48% were of high to very high risk [11].

While multiple options for reducing diarrheal prevalence by providing clean drinking water and using proper hand hygiene exist, inexpensive but potentially highly effective, point-of-use solutions remain an under-utilized option. One such option is the Sawyer^®^ PointONE™ water filter. Laboratory tests with the Sawyer^®^ PointONE™ water filter suggest it aligns with the United States Environmental Protection Agency standard for bacteria and protozoa removal [12]. Prior studies have identified a significant decrease in diarrhea prevalence [12, 13]. Some studies have argued that filters in the field have been fouled and under-utilized in practice [14, 15], however, others have noted numerous limitations of these studies [16] and reasonably good performance at removing *E. coli* and coliforms over a one- to three-year period in the field [17]. Thus, there is a continued need for carefully designed field trials to evaluate the efficacy of Sawyer® PointONE™ and other hollow fiber membrane filters.

In an attempt to better understand filter efficacy, utilization, and the impact of deployment in different community locations, we designed a prospective, randomized, controlled trial whereby 16 villages in the Dominican Republic were selected and randomly assigned to one of four treatment arms based on the location of Sawyer^®^ PointONE™ filter installation (filter in both home and school; filter in home only; filter in school only; control group). Village households were followed over time, monitoring self-reported health characteristics. Drinking water samples were also obtained from a subsample of households to monitor water quality for bacterial and chemical contamination in order to, first, directly assess filter removal of bacteria and particulates and, second, to assess whether the sole use of this type of filter as an intervention was contraindicated due to the presence of toxic levels of dissolved heavy metals.

## Methods

Sixteen villages in Dominican Republic were selected for inclusion in the study. Each selected village sent school-aged children to a private school within a pre-identified private school network. Each school draws its students from a unique geographic area (i.e., village) surrounding the school. Rural and urban villages across the country were included (Figure 1). The geographic distribution of schools (North, South, East, Capital-1, Capital −2 where “Capital” indicates schools in and near Santo Domingo) is shown in Table 1. Donor funding was available through a non-profit organization to provide point-of-use water filtration systems (Sawyer^®^ PointONE™, additional details on filter construction described elsewhere [13]) to in-network, sponsored households with a school-aged child that attended the private school. The total number of sponsored households eligible for inclusion at the start of the study in August 2018 was 675, with breakdown by village shown in Table 1.

**Table 1.** Household characteristics by village.

| De-identified village name | Total Sponsored Households Eligible for inclusion<sup>1</sup> | Total households in primary analyses<sup>2</sup> | Village Response Rate | Initial Treatment Group<sup>3</sup> | Geographic location | School received filter?<sup>4</sup> |
| --- | --- | --- | --- | --- | --- | --- |
| A | 26 | 25 | 96.2% | Control | North | No |
| B | 30 | 21 | 70.0% | Simultaneous | Capital – 1 | Yes |
| C | 35 | 17 | 48.6% | Control | Capital – 2 | No |
| D | 32 | 5 | 15.6% | Home | Capital – 1 | No |
| E | 46 | 38 | 82.6% | Control | North | No |
| F | 60 | 41 | 68.3% | Home | North | No |
| G | 18 | 12 | 66.7% | Home | North | No |
| H | 40 | 36 | 90.0% | School | Capital – 2 | Yes |
| I | 38 | 22 | 57.9% | Control | South | Yes |
| J | 24 | 3 | 12.5% | School | East | Yes |
| K | 72 | 4 | 5.6% | Home | East | No |
| L | 50 | 16 | 32.0% | Control | East | No |
| M | 47 | 32 | 68.1% | Home | Capital – 2 | Yes |
| N | 62 | 8 | 12.9% | Control | East | Yes |
| O | 50 | 15 | 30.0% | Simultaneous | South | Yes |
| P | 45 | 27 | 60.0% | Home | Capital – 1 | No |
| Total | 675 | 322 | 47.7% | -- | -- | -- |

<sup>1</sup>Number of sponsored households within the village as of August 2018.
<sup>2</sup>Use in primary analyses means that at least one household survey pre-household filter installation and at least one household survey post-household filter installation are available, consent of the respondent was received and at least 90% of the survey question responses were valid/not missing.
<sup>3</sup>Since all households and eligible schools ultimately received a filter, treatment groups listed in the table are “Initial” groups with subsequent installation of school/home measurements and commensurately collected survey data.
<sup>4</sup>Nine schools declined receiving a filter at the school because they determined they were not in need of a filter.
Note: No water samples were taken before this determination was made.

**Figure 1.**
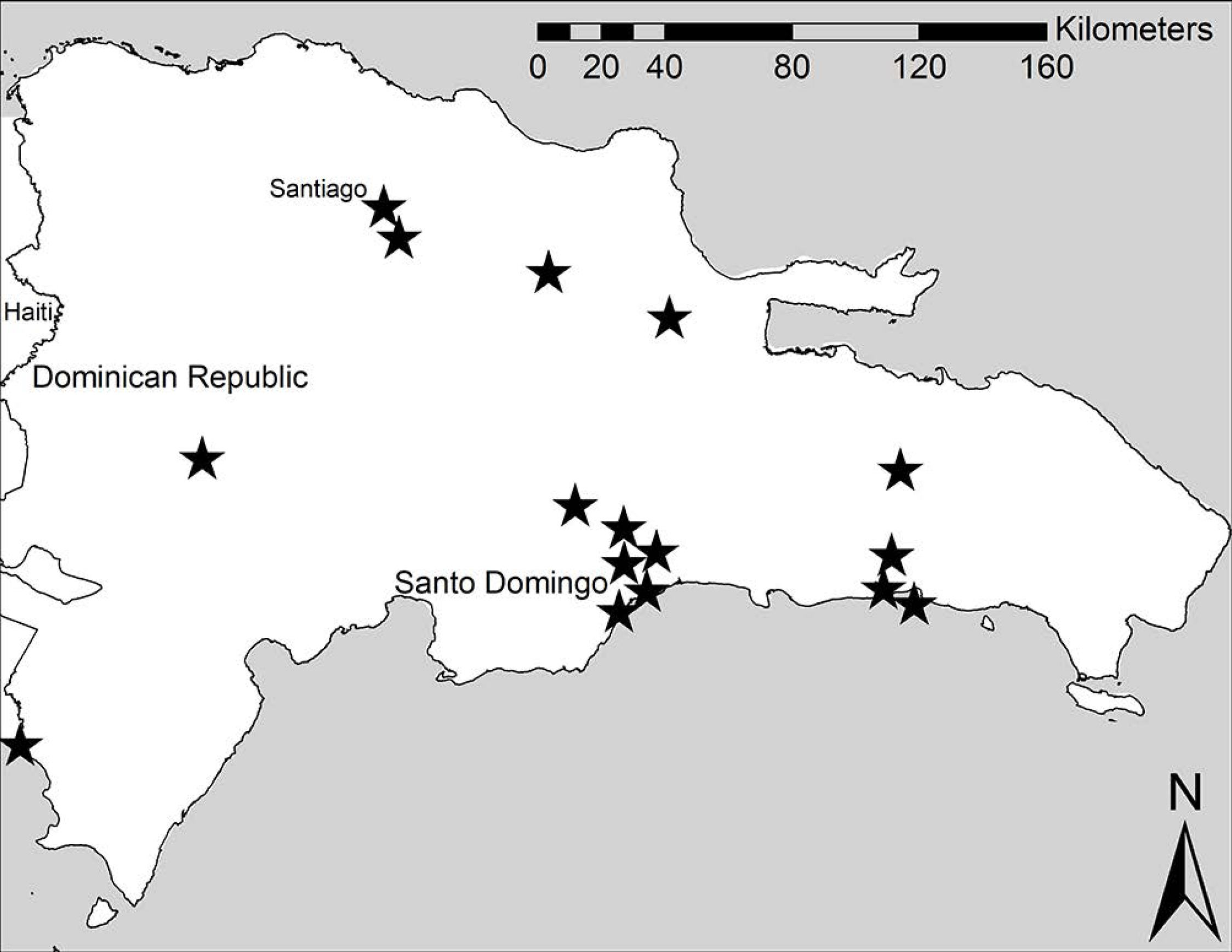
Location of 16 selected villages in the Dominican Republic. Stars denote the approximate location of villages included in this study. Villages that were close in proximity were offset on the map to provide distinct representations for each village.

### Randomization procedures

In order to evaluate the efficacy of filter installations in both schools and homes, a modified factorial design was used (Figure 2). Each of the sixteen villages was initially assigned to one of the following treatment groups: control (no filter initially), home filter only, school filter only or simultaneous (school and home) filter installation. Assignment of villages to treatment groups (see Table 1) followed a covariate adaptive randomization strategy, whereby predefined covariates are balanced across treatments using the method of minimization [18]. We performed the method of minimization including the following covariates: (a) reported number of sponsored children at each school as of August 2018 (high [51+]) vs. low [50 or fewer]), (b) geographic location/watershed (five groups; [19]), (c) whether the school anticipated receiving a filter or not (some schools declined needing filters due to already having a self-reported ‘safe drinking water solution’) and (d) government public health statistics regarding endemic levels of diarrhea in the region [9] in children under 5 (15% or more vs. 15% or less). Villages initially assigned to the control group subsequently were assigned to the school filter or home filter treatment group. All treatment groups received filters in the home by the end of the study.

**Figure 2.**
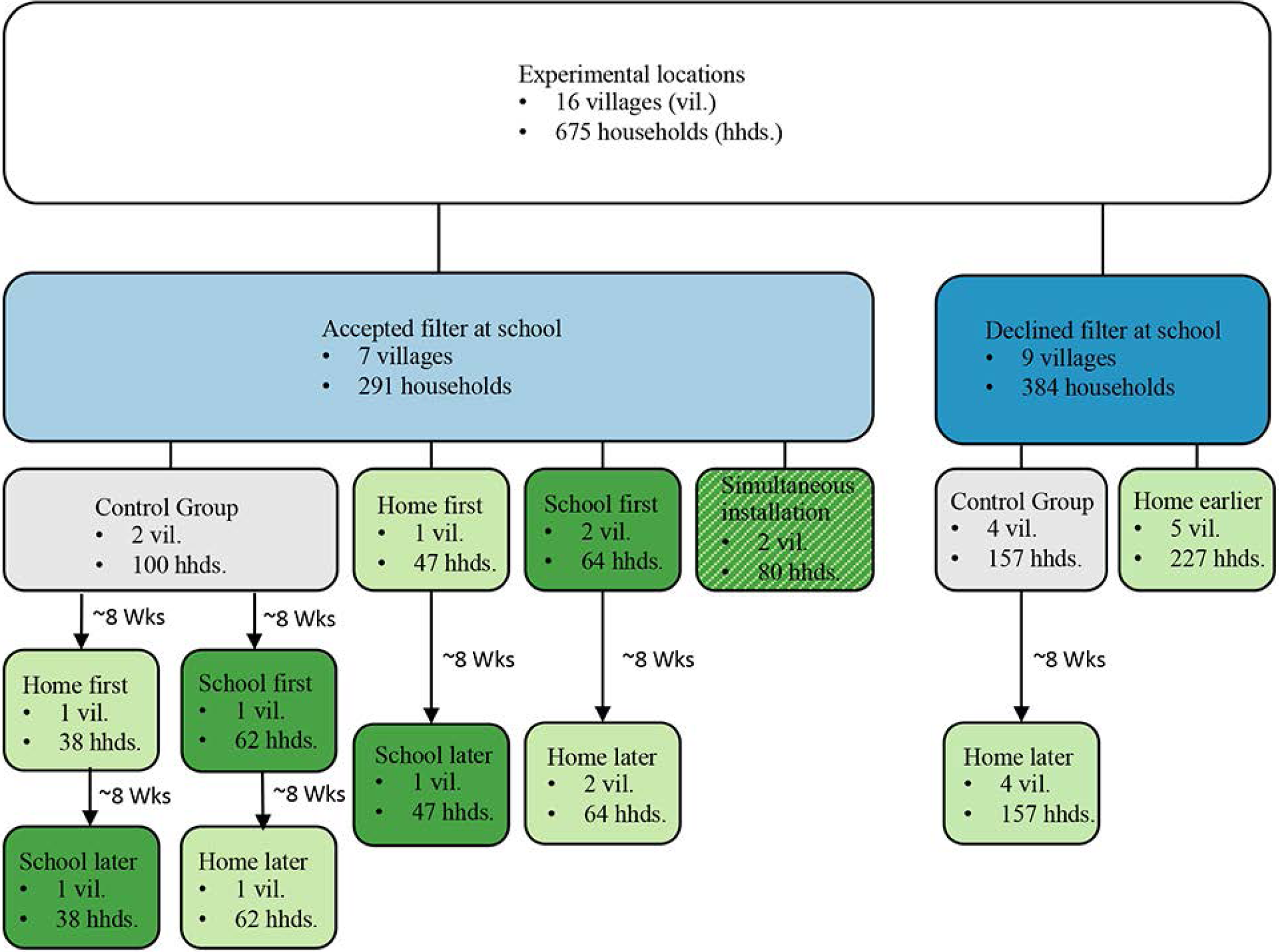
Depiction of the assignment of schools and households to treatment groups. Depending upon whether the school in the village accepted or declined a filter, villages were initially assigned to either four or two treatment groups, respectively. In order to ensure that all households and schools that accepted a filter ultimately received one, secondary randomization of control group households was performed as depicted in the figure.

### Survey and data collection

We administered a short demographic, health and economic baseline questionnaire to all households upon installation of point-of-use filter in the home [13]. Five local data collectors were trained by the research team. Each data collector was assigned to a village based on pre-determined geographic areas (Table 1) and was the same person for pre-filter (control group; 2-8 weeks prior to filter installation), baseline (filter installation) and follow-up surveys at respective households and schools. Data collection took place from September 2018 through April 2019.

Filter installation included a brief training on use of the filter by the data collector, mention of importance of handwashing and a demonstration by the respondent of proper filter use and backflushing. After installation of the filter, data collectors attempted to collect two-, eight- and sixteen-week follow-up surveys. This study was approved and monitored by Dordt University IRB and all surveys included a statement of consent for research use. The trial is registered with www.clinicaltrials.gov as NCT03972618.

Eligibility for inclusion in analyses in this manuscript required: (a) at least one survey at or before the time of filter installation (pre-survey), (b) at least one survey completed at least two weeks after filter installation (post-survey), (c) obtained consent for data to be used for research purposes and (d) having provided answers to at least 90% of the survey questions. Using these criteria, the overall eligible data response rate was 47.7% (322/675 households; see Table 1) with variation in response rates by village (minimum = 5.6%; maximum = 96.2%) and region (generally lower response rates in the lowlands (East) region).

Across the 322 households in the primary analysis, there were 1075 household-level survey administrations. These household surveys consisted of 72 control group baseline survey responses, 322 baseline survey responses upon filter installation, and 681 follow-up survey responses (mean number of follow-up survey responses per household was 2.11). Among the 681 follow-up survey responses, 107 of the responses indicated not using the filter and thus were ignored in primary analyses. An intent to treat sensitivity analysis included these households.

### Survey variables

The primary outcome we considered was self-reported, two-week prevalence of diarrhea, including whether diarrhea had economic or educational consequences, causing hospitalization and/or missed school or work. The household survey administration included questions that determine: (a) the size of the household (number of adults and children living in the household), and then for each household member, whether they have in the past two weeks, (b) had diarrhea, and if so, whether that diarrhea (c) caused missed days of work (adults) or missed days of school (children), and (d) required hospitalization. Additional information used in analysis was (1) season (Fall [Oct, Nov], Winter [Dec, Jan, Feb] or Spring [Mar, Apr]), (2) days since filter installation, (3) region (East, West, South, Central – 1, Central-2), and (4) water source (city, purchased, well or other (e.g. river, catchment)).

### Water sampling methods

Between two and seven untreated drinking water source samples were collected at each of the sixteen locations (56 total samples). Information on location, date of the sample acquisition and whether the sample came from a home or from the school can be found in Additional File 1. Water samples were tested for the presence of dissolved heavy metals, heavy metals associated with particulate matter, and potential bacterial pathogens using Sawyer^®^ PointONE™ hollow fiber membrane filters to capture particulates and bacteria and using a metal chelating foam to capture dissolved heavy metals. ICP-OES was used to measure dissolved heavy metals that have been identified by the WHO as potential health hazards (Arsenic, Barium, Cadmium, Chromium, Copper, Lead, Nickel and Selenium) [20]. A combination of spectrophotometry, SEM-EDS, and powder X-Ray diffraction (PXRD) were used to determine the concentration of particulates (mg/L) and whether those particulates contained heavy metals of interest. Lastly, the presence of eighteen bacterial genera identified by the WHO as containing potential waterborne pathogens [21] (*Acinetobacter, Aeromonas, Burkholderia, Campylobacter, Enterobacter, Escherichia*/*Shigella, Francisella, Helicobacter, Klebsiella, Legionella, Leptosipra, Mycobacterium, Pseudomonas, Salmonella, Staphylococcus, Tsukamurella, Vibrio* and *Yersinia*) were assessed through 16S rRNA amplicon sequencing following the Schloss Lab MiSeq SOP [22, 23] for sequencing and initial data processing to produce 97% operational taxonomic units and assigned taxonomies using the Silva Release 132 alignment and database [24]. Downstream analyses of 16S amplicon sequencing data were performed using the *R* packages Phyloseq [25] and vegan [26]. Details of water sampling, testing and analysis methods are available in Additional File 2. Sequencing data associated with this study have been deposited in the Short Read Archive under PRJNA670359 (https://www.ncbi.nlm.nih.gov/bioproject/PRJNA670359).

### Statistical analysis

Generalized linear mixed-effects models with a logistic link function were used to predict diarrhea prevalence by filter status. Random effects were used for repeated measures, with additional fixed effects covariate adjustments for season, water source and household size. Generalized linear mixed effects models with random effects for repeated measures were also used to compare water sampling data between pre- and post-filter installation measurements. All statistical analyses were completed using *R* version 3.6.2 [27] and used a significance level of 0.05.

## Results

### Characteristics of the villages

Characteristics of the villages are shown in Table 2. Purchased water usage fell from 36% to 10% from baseline to follow-up, likely reflecting reliance on the filter to provide safe and clean water. Modest, though statistically significant, changes in region were observed from baseline to follow-up (e.g., Capital – 1 accounted for 21% of follow-up data, but only 12% of baseline data due to proportionally more follow-up surveys conducted in this region as compared to others). As expected, the percent of households with a filter at their children’s school was somewhat higher at follow-up (31% vs. 18%).

**Table 2.** Village Characteristics.

| Variable | Before home filter<br>installation (n=394) | After home filter<br>installation (n=574) | P-value for<br>difference<sup>1</sup> |
| --- | --- | --- | --- |
|  | Mean (SD) or<br>%(x/n) | Mean (SD) or<br>%(x/n) |  |
| Household size | 4.60 (1.72) | 4.41 (1.33) | 0.049* |
| Region |  |  |  |
| North | 38.8% (153/394) | 35.0% (201/574) | 0.004** |
| East | 8.6% (34/394) | 5.9% (34/574) |  |
| South | 15.0% (59/394) | 15.5% (89/574) |  |
| Capital – 1 | 12.4% (49/394) | 21.4% (123/574) |  |
| Capital – 2 | 25.1% (99/394) | 22.1% (127/574) |  |
| School filter status |  |  |  |
| Before/Declined | 82.0% (323/394) | 68.6% (414/594) | <0.001*** |
| After | 18.0% (71/394) | 31.4% (180/594) |  |
| Season |  |  |  |
| Fall | 67.3% (265/394) | 19.2% (110/574) | <0.001*** |
| Winter | 32.7% (129/394) | 57.3% (329/574) |  |
| Spring | 0.0% (0/394) | 23.5% (135/574) |  |
| Water source |  |  |  |
| City | 49.4% (195/394) | 79.6% (457/574) | <0.001*** |
| Purchased | 36.0% (142/394) | 10.3% (59/574) |  |
| Well | 10.4% (41/394) | 8.5% (49/574) |  |
| Other | 4.1% (16/394) | 1.6% (9/574) |  |

\* $p < 0.05$ ; \*\* $p < 0.01$ ; \*\*\* $p < 0.00$
<sup>1</sup> Logistic regression model with random effects term accounting for repeated measures.

### Two-week diarrhea prevalence by home filter status

Before filter installation, 25.6% of households reported at least one member of the household experiencing diarrhea within the previous two weeks (Table 3; Figure 3). Prevalence of diarrhea dropped substantially to 9.8% after filter installation, a difference which remained statistically significant even after adjusting for other variables (adjusted Odds Ratio (aOR) =0.29 (95% CI: 0.16, 0.51), p<0.001). Adjusted odds ratios were statistically significant for adult-only diarrhea and for children under 4 years (0.14 and 0.09, respectively). While the aOR was still small (0.49) it was not statistically significant (p=0.07) for school-aged children after adjusting for other variables (Table 3). We also completed an intent to treat analysis including the 107 households reporting not using the filters (total n=681) which demonstrated higher post filter installation diarrhea prevalence overall (10.3%), as well as in Adults (4.8%) and Children under the age of 5 (7.1%, with slightly lower prevalence in school aged children (5.7%; Additional File 3).

**Table 3.**
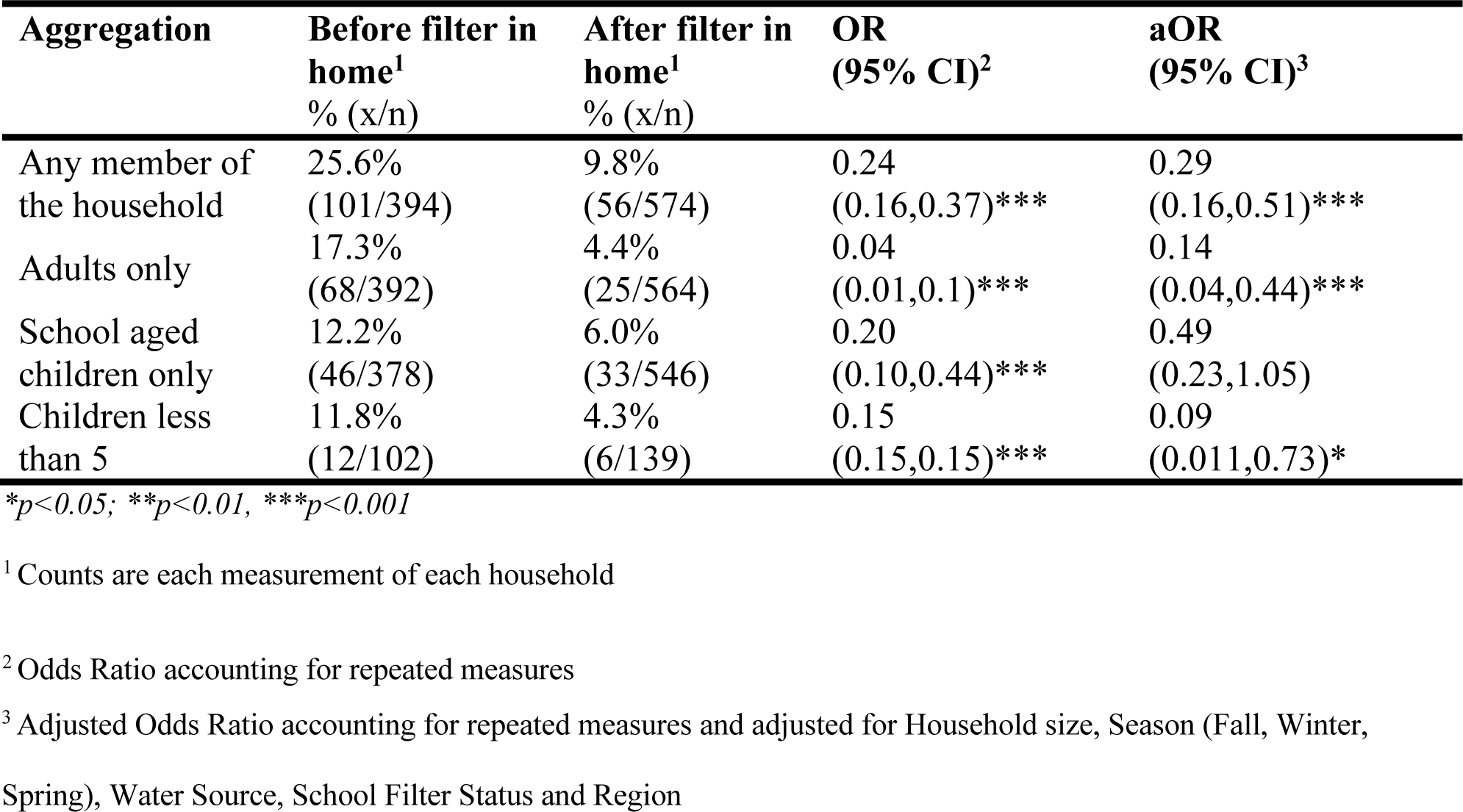
Prevalence of self-reported diarrhea by home filter status.

**Figure 3:**
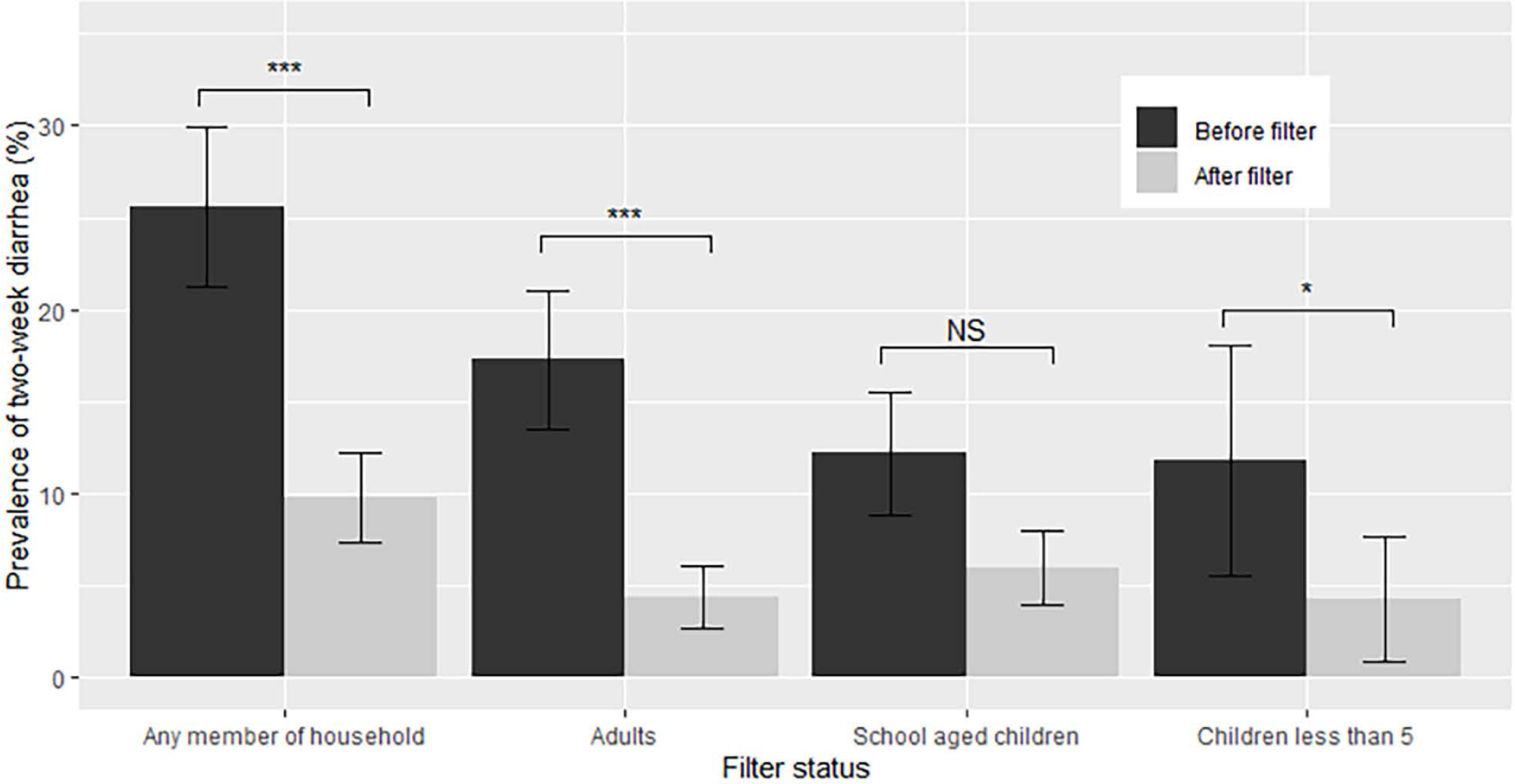
Prevalence of two-week diarrhea before and after filter installation by age. Unadjusted SE bars are shown. Statistical significance between groups (***p<0.001, **p<0.01, *p<0.05; NS p>0.05) represents statistical significance from repeated measures models adjusted for Household size, Season (Fall, Winter, Spring), Water Source, School Filter Status and Region

**Figure 4.**
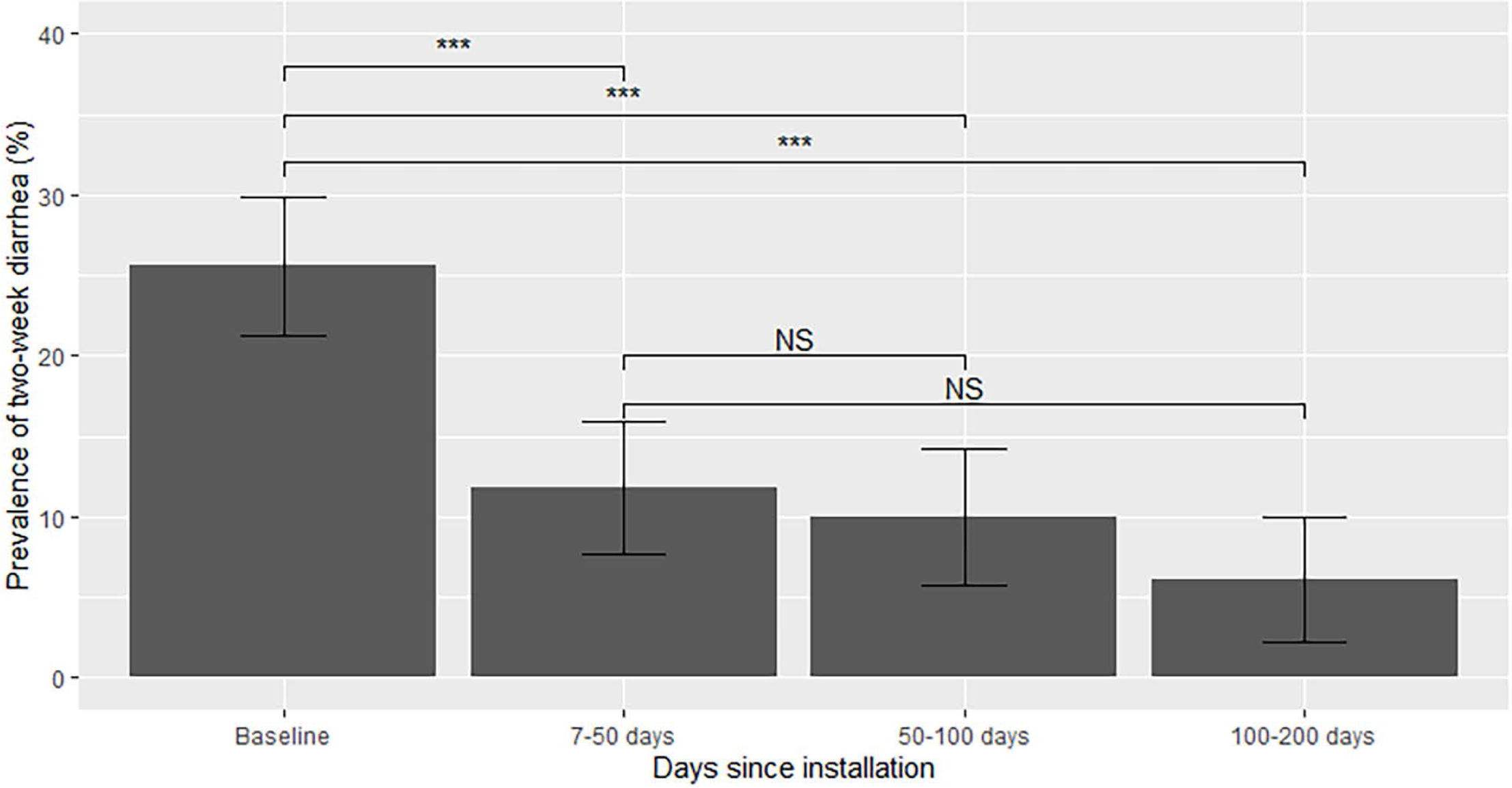
Diarrhea prevalence by days since installation. Unadjusted SE bars are shown. Statistical significance between groups (***p<0.001, **p<0.01, *p<0.05; NS p>0.05) represents statistical significance from repeated measures models adjusted for Household size, Season (Fall, Winter, Spring), Water Source, School Filter Status and Region

**Figure 5.**
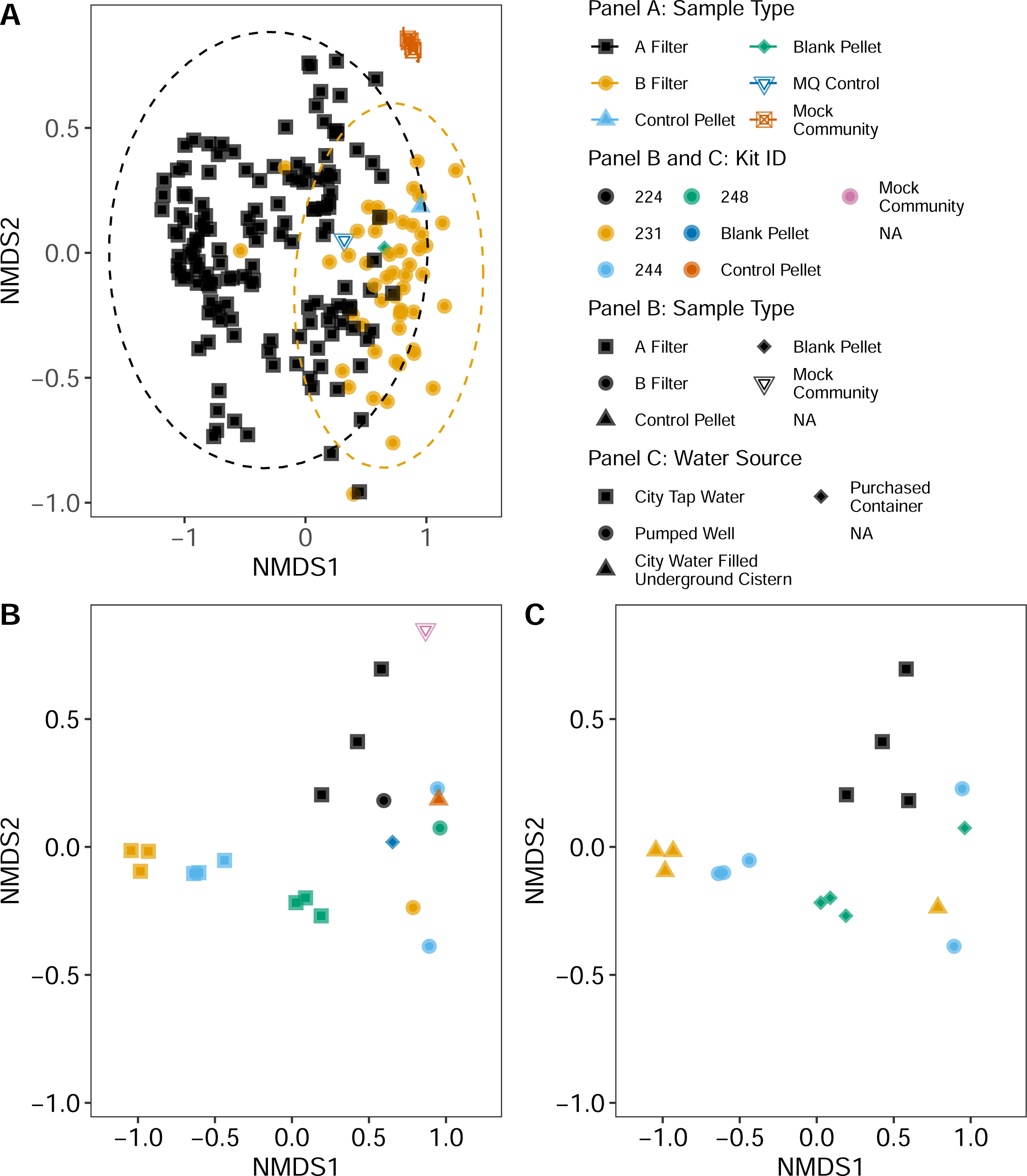
NMDS ordination (k=3, stress = 0.141) of Bray-Curtis pairwise distances of bacterial communities from drinking water sources collected in the study (either home or school locations). Each water source was sampled with a kit consisting of three tandem filters (Filter A and Filter B), yielding three biological replicates of a water source per kit. Each point represents backflushed contents from Filter A, Filter B, laboratory backflush controls, mock community controls, or negative controls. Each kit is represented by up to 3 Filter A points and 3 Filter B points. Samples with fewer than 5000 sequencing reads were not included in the analysis. The same ordination is used for all three panels. Panel A: All backflush samples included in the study and associated controls. Shape and color delineate whether a sample is from Filter A, Filter B or a control. Panel B: A subset of four kits selected to show the relationship between biological replicates of Filter A and Filter B within and between kits. Shape delineates Filter A, Filter B and control samples. Color delineates which kit (i.e. water source) each filter sample represents along with the type of control. Panel C: The same kits as shown in Panel B with identical coloring showing kit membership. Shape delineates the type of water source.

Prevalence of diarrhea was significantly lower than baseline throughout this study up to 200 days post filter installation (adjusted p<0.001 for all three of: = 7-50 days vs. baseline; 50-100 days vs. baseline and 100-200 days vs. baseline). While diarrhea prevalence continued to decline over time (11.8% 7-50 days; 10.0% 50-100 days; 6.1% 100-200 days), these differences were not statistically significant after accounting for repeated measures, seasonal and regional differences (7-50 days vs. 50-100 days (adjusted p=0.45); 7-50 days vs. 100-200 days (adjusted p=0.38)).

### Two-week diarrhea prevalence by school filter status

We also examined whether two-week prevalence of diarrhea was impacted by the installation of water filters in schools, irrespective of water filters in the home. Before or after adjusting for covariates, there was no evidence of statistical interaction between school filter status and home filter status for any household member, adults, school-aged children, or children less than 5 (Adjusted p-values of 0.63, 0.70, 0.76 and 0.99, respectively). Furthermore, there was no evidence of a statistically significant decline in two-week diarrhea after installation of school filters, for any age group (Table 4) before or after adjusting for covariates, including home filter status.

**Table 4.** Prevalence of self-reported diarrhea in household by school filter status.

| Aggregation | Before filter in school<sup>1</sup><br>% (x/n) | After filter in school<sup>1</sup><br>% (x/n) | OR<br>(95% CI)<sup>2</sup> | aOR<br>(95% CI)<sup>3</sup> |
| --- | --- | --- | --- | --- |
| Any member of the household | 15.9% (114/717) | 17.1% (43/251) | 1.01 (0.62,1.65) | 1.29 (0.69,2.40) |
| Adults only | 9.62% (68/707) | 10% (25/249) | 0.87 (0.40,1.88) | 1.58 (0.34, 7.39) |
| School aged children only | 7.99% (55/688) | 10.2% (24/236) | 1.09 (0.51,2.33) | 1.17 (0.55,2.52) |
| Children less than 5 | 7.18% (13/181) | 8.33% (5/60) | 0.34 (0.018,6.33) | 0.47 (0.09,2.54) |

\*p<0.05, \*\*p<0.01, \*\*\*p<0.001
<sup>1</sup> Counts are each measurement of each household
<sup>2</sup> Odds Ratio accounting for repeated measures
<sup>3</sup> Adjusted Odds Ratio accounting for repeated measures and adjusted for Household Filter presence (y/n), Household size, Season (Fall, Winter, Spring), Water Source and Region

### Economic and educational consequences of diarrhea

Prevalence of diarrhea with economic or educational consequences decreased significantly across all age groups, and for each consequence (i.e., missed work, missed school, hospitalizations) across all age groups (all p<0.05 before adjusting for covariates) after filters were installed in homes (Table 5). Before filters were installed in homes, prevalence of diarrhea with economic or educational consequences was 9.64% among any member of the household, compared to 1.57% after filter installation (p<0.001). For children, missed school due to diarrhea decreased from 4.23% to 0.55% (p<0.001). After adjusting for covariates (Table 5), prevalence of diarrhea with any economic or educational consequence for any member of the household still showed a statistically significant decrease after filter installation (aOR 0.07, p<0.05), while other decreases were no longer statistically significant. There was no evidence of an impact of the school filter or interaction between school and home filters on diarrhea with economic or educational consequences (p>0.05 in all cases; detailed results not shown).

**Table 5.** Prevalence of diarrhea with economic or educational consequence by home filter status.

| Aggregation | Before filter in home<sup>1</sup> | After filter in home<sup>1</sup> | Odds ratio<sup>2</sup> | Adjusted odds ratio<sup>3</sup> |
| --- | --- | --- | --- | --- |
| Any member of the household with any economic or educational consequence | 9.64% (38/394) | 1.57% (9/574) | 0.01 (0.00,0.05)*** | 0.07 (0.0014,0.56)* |
| Adult hospitalized due to diarrhea | 3.59% (14/390) | 0.888% (5/563) | 0.02 (0.00,0.18)*** | 0.18 (0.0027,11.6) |
| Adult missed work due to diarrhea | 5.38% (21/390) | 0.888% (5/563) | 0.01 (0.00,0.09)*** | 0.02 (0.00,1.57) |
| Child missed school due to diarrhea | 4.23% (16/378) | 0.549% (3/546) | 0.01 (0.00,0.09)*** | 0.051 (0.0016,1.59) |
| Child less than 5 hospitalized due to diarrhea | 7.84% (8/102) | 2.88% (4/139) | 0.01 (0.00,0.32)* | 0.01 (0.00,12.0) |

\*p<0.05, \*\*p<0.01, \*\*\*p<0.001
<sup>1</sup> Counts are each measurement of each household
<sup>2</sup> Odds Ratio accounting for repeated measures
<sup>3</sup> Adjusted odds ratio accounting for repeated measures and adjusted for Household size, Season (Fall, Winter, Spring), Water Source, School Filter Status and Region

**Table 5.**
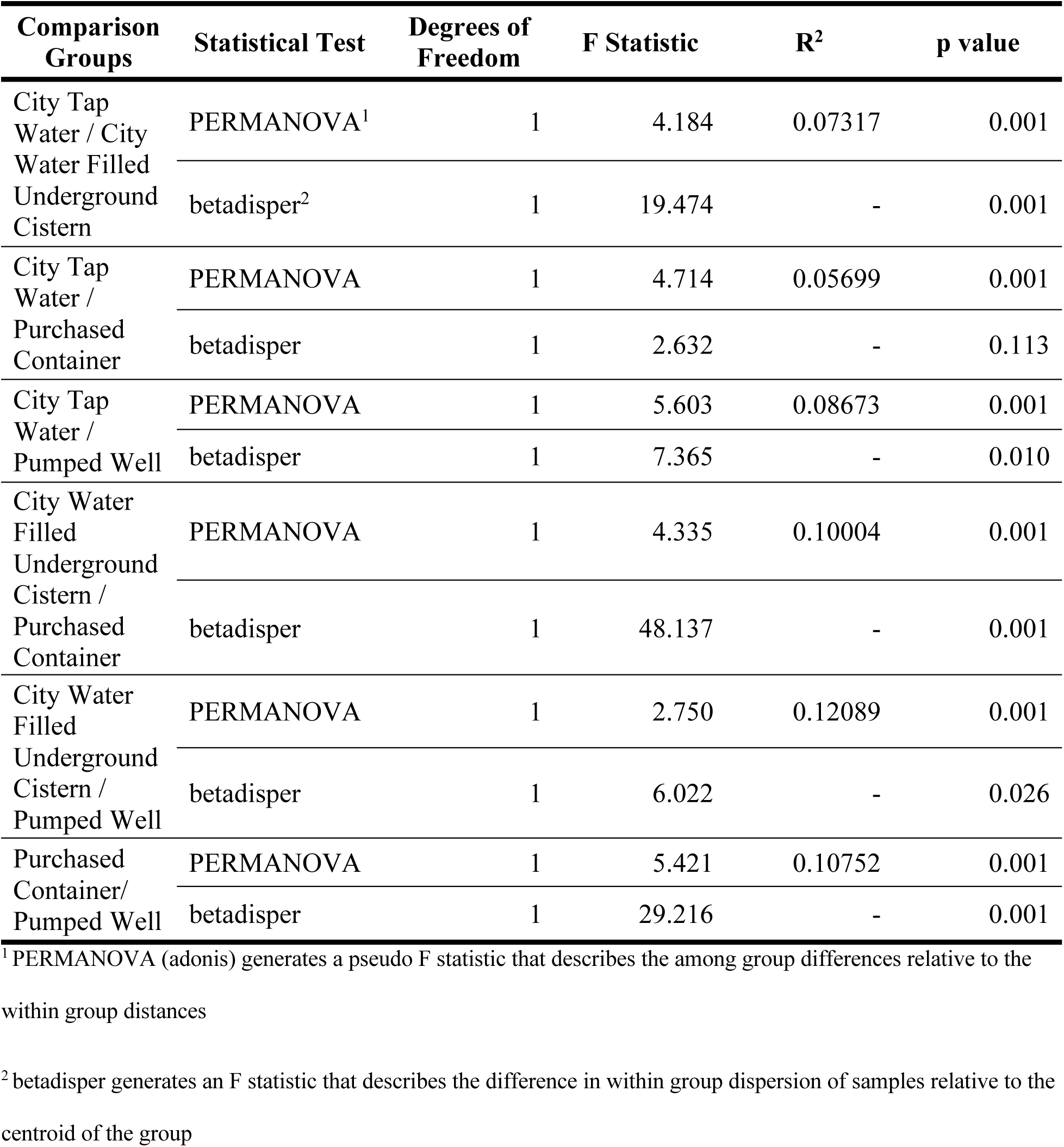
Statistical Tests of Bacterial Communities by Water Source Type.

### Quality of drinking water sources

In order to assess the quality of drinking water sources, 56 unfiltered water samples were collected across the 16 villages (30 home water sources, 25 school water sources, 1 unknown) throughout the course of the study. Thirty-five samples were collected at the time of filter installation and twenty-one after filter installation. Characteristics of each water sample and outcomes for detection of dissolved heavy metals, particulates and bacteria are provided in Additional File 1.

We analyzed bacterial communities of drinking water sources through 16S rRNA amplicon sequencing. Up to 5 gallons (∼19 L) of water from each drinking source was filtered through two, 0.1 micron hollow fiber membrane filters linked in a tandem pair. The first filter in the pair (designated “Filter A”) filtered the source water, capturing cells and particulate matter; the second filter in the pair (designated “Filter B”) captured cells and particulate matter (if any) of the filtrate of Filter A. This design allowed the analysis of bacterial communities of both the source water and the filtered water. Two types of controls were also analyzed. First, on each day of processing, backflush controls included 1) water backflushed through an unused filter, 2) a full volume of water used for backflushing all filters was pelleted, and 3) a small volume of water used for backflushing all filters was added to DNA extraction tubes. Second, each sequencing library plate included a positive control of a mock community of 8 bacteria (ZymoBIOMICS Microbial Community Standard) and a negative control consisting of ultrapure water. Evaluation of the pairwise distances between bacterial communities of each filter in the study (Fig 6A) showed that the communities derived from unfiltered source water (Filter A) formed a group that is largely non-overlapping with the communities derived from filtered source water (Filter B). The difference between the two groups was statistically significant (permutational multivariate analysis of variance [PERMANOVA] [28]; df = 1, F = 9.3361, R^2^ = 0.04755, p = 0.001), however, within group dispersion may also contribute to between group significance (betadisper, F = 4.1054, p = 0.045). Biological replicates of each water source grouped tightly in most cases for the unfiltered water (Filter A), whereas the replicates for filtered water (Filter B) from the same kit did not cluster as tightly (Fig. 6B). Replicate Filter B samples are generally co-localized with Filter B samples from other kits and with backflush negative controls (Fig. 6B). Consistent with the patterns in Fig. 6B, Filter A replicates from the same kit were less distant from each other than Filter A communities from other kits, explaining a large fraction of the observed variance (PERMANOVA; Filter A: df = 54, F = 7.3144, R^2^ = 0.82808, p = 0.001; betadisper, F = 2.8424, p = 0.002). Similar patterns were observed for Filter B communities (Filter B: df = 32, F = 1.9172, R^2^ = 0.76354, p = 0.001; betadisper, F = 27.287, p = 0.005). There were distinct bacterial communities associated with samples from different types of water sources. For example, each of the kits in Fig. 6C represents a different water source type and is representative of the positions in the ordination of other kits of the same water source type sampled from different geographic locations. Each of the water type bacterial communities are distinct from the other water type bacterial communities in pairwise comparisons of all samples, with all but one comparison having significant differences both between (PERMANOVA) and within (betadisper) group distances (Table 5).

We used the list of bacterial taxa identified in each water source to determine the presence or absence of genera containing known waterborne pathogens as defined by the WHO [21]. A genus was designated as present if it was found in A filter replicates for a kit but not found in B filters or process controls. At least one genus from the list of 18 waterborne bacterial pathogens was detected in 50 of 51 water sources passing quality control metrics. The single water source without indication of potential pathogens was from a city tap water sample (Kit 233). Additionally, we tested for the presence/absence of heavy metals in water sources. We detected at least one of eight dissolved heavy metals in 38 out of 56 water samples (67.9%). Table 6 shows detected bacterial pathogens and dissolved heavy metals stratified by whether the sample was obtained at the time of filter installation or during a follow-up visit. No significant differences were observed in the prevalence of detected bacteria or heavy metals in the source water between installation and follow-up across all measured bacteria and heavy metals measured. Similarly, no seasonal or linear changes in prevalence were observed over time. Furthermore, mean particulate levels were not significantly different at the time of filter installation (0.246 vs. 0.166 mg/L; p=0.57), and also did not differ significantly over time during the study (linear trend p=0.36; seasonal effect p=0.35). Detected particulates were found to contain Ba, Ce, Cl, Cr, Fe, P, Mn, S, Ti, Zr. See Additional File 1 for details about all water quality results.

**Table 6.** Summary of test results of source water (unfiltered)

| Type of water sample testing | Bacterial Genus /Heavy Metal | Percent of samples present at filter install | Percent of samples present at follow-up | p-value comparing install to follow-up<sup>2</sup> | P-value for linear trend<sup>3</sup> | P-value for seasonal effect<sup>4</sup> |
| --- | --- | --- | --- | --- | --- | --- |
| **Bacteria detected** | *Acinetobacter* | 53.8% (14/26) | 35.3% (6/17) | 0.25 | 0.39 | 0.56 |
|  | *Aeromonas* | 16.1% (5/31) | 20% (4/20) | 0.72 | 0.51 | 0.64 |
|  | *Burkholderia* | 6.45% (2/31) | 5% (1/20) | 0.83 | 0.31 | 0.54 |
|  | *Campylobacter* | 0% (0/34) | 0% (0/22) | 1.00 | 1.00 | 1.00 |
|  | *Enterobacter* | 0% (0/34) | 0% (0/22) | 1.00 | 1.00 | 1.00 |
|  | *Escherichia/Shigella* | 45.5% (10/22) | 50% (6/12) | 0.80 | 0.90 | 0.90 |
|  | *Francisella* | 0% (0/34) | 0% (0/22) | 1.00 | 1.00 | 1.00 |
|  | *Helicobacter* | 0% (0/34) | 0% (0/22) | 1.00 | 1.00 | 1.00 |
|  | *Klebsiella* | 0% (0/34) | 0% (0/22) | 1.00 | 1.00 | 1.00 |
|  | *Legionella* | 58.1% (18/31) | 70% (14/20) | 0.18 | 0.06 | 0.41 |
|  | *Leptospira* | 0% (0/34) | 0% (0/22) | 1.00 | 1.00 | 1.00 |
|  | *Mycobacterium* | 30% (9/30) | 57.9% (11/19) | 0.00 | 0.19 | 0.08 |
|  | *Pseudomonas* | 85.2% (23/27) | 94.4% (17/18) | 0.35 | 0.62 | 0.40 |
|  | *Salmonella* | 0% (0/34) | 0% (0/22) | 1.00 | 1.00 | 1.00 |
|  | *Staphylococcus* | 12.9% (4/31) | 15.8% (3/19) | 0.78 | 0.79 | 0.87 |
|  | *Tsukamurella* | 0% (0/34) | 0% (0/22) | 1.00 | 1.00 | 1.00 |
|  | *Vibrio* | 0% (0/34) | 0% (0/22) | 1.00 | 1.00 | 1.00 |
|  | *Yersinia* | 3.23% (1/31) | 0% (0/20) | 0.98 | 0.69 | 0.47 |
| **Dissolved heavy metal ions detected** | Arsenic | 0% (0/34) | 0% (0/22) | 1.00 | 1.00 | 1.00 |
|  | Barium | 0% (0/34) | 0% (0/22) | 1.00 | 1.00 | 1.00 |
|  | Cadmium | 0% (0/34) | 0% (0/22) | 1.00 | 1.00 | 1.00 |
|  | Chromium | 0% (0/34) | 0% (0/22) | 1.00 | 1.00 | 1.00 |
|  | Copper<sup>1</sup> | 64.7% (22/34) | 68.2% (15/22) | 0.79 | 0.38 | 0.34 |
|  | Lead<sup>1</sup> | 8.82% (3/34) | 4.55% (1/22) | 0.55 | 0.38 | 0.63 |
|  | Nickel | 0% (0/34) | 0% (0/22) | 1.00 | 1.00 | 1.00 |
|  | Selenium<sup>1</sup> | 5.88% (2/34) | 0% (0/22) | 0.96 | 0.16 | 0.21 |

<sup>1</sup> Detected levels were below WHO action guidelines in all samples
<sup>2</sup> From Fisher's exact test comparing baseline to follow-up prevalence
<sup>3</sup> From test of linear trend over course of study
<sup>4</sup> From test of seasonal differences over course of study

## Discussion

The purpose of this study was to evaluate the impact of the introduction of point-of-use filters into different community settings on the health of school aged children and their families. Drinking water sources were also evaluated for bacterial, particulate and dissolved heavy metal content to determine the appropriateness of the point-of-use filter technology for long term use by residents. Two-week self-reported diarrhea prevalence in all age groups drops substantially after introduction of filters in homes, with unadjusted declines of between approximately 50 and 75%. Similarly, large reductions in economic and educational consequences of diarrhea were also observed after introduction of filters in homes across all age groups. No corresponding change in prevalence was noted after introduction of filters into schools, nor was any moderating effect found for the school filter on the effectiveness of the home filter. Results from water sample testing confirm that bacteria were prevalent in water samples, including genera that are known to contain pathogens associated with waterborne disease. Limited amounts of dissolved metals and metal-containing particulates were observed in the drinking water sources.

While previous research [13–15, 17] yielded mixed results from the Sawyer^®^ PointONE™ filter system in laboratory testing and field environments, our randomized, controlled field trial provides additional evidence of their effectiveness in reducing waterborne disease out to at least 200 days post-installation in real world settings. The hollow fiber membrane technology is designed to remove bacteria and particulates, but not dissolved ions. Evaluation of unfiltered drinking water sources available to families confirmed the presence of highly diverse bacterial communities that varied with source type. Only a single water source tested in this study was free of bacterial genera that are known to contain waterborne pathogens, indicating that hollowfiber membrane filtration should be an effective method for improving drinking water quality and health outcomes in these areas. None of the drinking water sources contained dissolved heavy metals above WHO action standards, suggesting that point-of-use filters were an appropriate solution for the villages and schools targeted by this intervention. Furthermore, prevalence of reported diarrhea dropped substantially, though was not eliminated completely, after filter installation in line with expectations that the filter eliminates bacterial and parasitic causes of diarrhea from water while not impacting other diarrheal causes (e.g., viral). However, despite these positives, further field work is needed, especially evaluation of filter effectiveness and utilization over longer time periods.

Interestingly, little impact of school filter installation on diarrhea prevalence was observed. There are likely numerous causes for this finding. First, and importantly, only school-aged children in the family receive water at the school and, thus, any potential direct impact of school filters would only be observed on school-aged children. However, in our data, this direct impact was not observed. Secondly, follow-up conversations with school and non-profit workers (personal communication, C. Nelson), suggest that, in part due to a relatively short school day (4 hours), children typically bring water from home or buy water (not necessarily clean and safe) from vendors near the school instead of getting water from school. Third, only seven of the sixteen schools in the study were willing to accept a filter for their school, with nine schools having determined that their water was of sufficient quality that no filter was needed, despite no *a priori* water testing.

An important feature of our study evaluating the potential effectiveness of both school and home filters was a sequential study design implementing the method of minimization to balance covariates. As has been noted by others [18] this method is rarely implemented in practice but can be highly effective. In our case, the method of minimization maximized statistical power by limiting the impact of potential confounding variables over time and ensuring a powerful study design. However, even here, there are additional variables that could have minimized the impact of covariates even further (e.g., proportion of households using unfiltered water in each community; proportion lacking sanitation services per region, etc.); these remain potentially confounding variables in our analysis. This, combined with the use of a self-report diarrhea measurement, represent two areas for future work and limitations of this study.

Another limitation of our study worth noting are the widely varying response rates by village. While the impact on our findings is minimal, due in part to the study design, additional statistical power and reduced impacts of potential confounding would be realized through consistently higher response rates across all villages. Some variation and lower response rates are expected when working in remote villages in countries and areas with limited infrastructure, however, having additional community buy-in and support (see previous paragraphs) may serve not only to improve actual intervention efficacy but also to improve statistical aspects of study design leading to more quantifiable and robust findings.

This study also combined public health surveys with evaluation of both bacterial and chemical components of drinking water quality using the same hollowfiber membrane filtration technology as was used for the intervention in homes and schools. The tandem filtration design of test kits provided a direct demonstration of removal of potentially harmful bacteria and particulate matter that could be associated with toxic metals. Dissolved heavy metals were assessed from the filtered water sources to determine if potential toxins might persist in the drinking water source. Efforts to evaluate drinking water sources should take into account the biological and chemical components of the water to ensure that proper intervention technologies are used.

It is important to note that this study produced profiles of the bacterial communities in each water sample through sequencing of 16S rRNA amplicons. The use of this sequencing-based evaluation of the presence/absence and relative abundance of bacterial genera is not traditionally applied in the evaluation of drinking water quality, although this is beginning to change [29–31]. More routinely, culture-based methods are used to assess viable loads of fecal indicator bacteria as a proxy for the presence of pathogens, or qPCR-based molecular methods are used to detect the presence of specific pathogens. When interpreting results from the sequencing approach employed in this study, it is important to recognize that the method does not determine whether bacteria detected in the sample are viable or not (this is also true of qPCR approaches). The method also does not identify specific pathogens (this is also true of routine culture-based approaches). However, it 1) allows for a single assay that characterizes the entire bacterial community, including all known potential pathogenic groups of bacteria, and 2) it provides an opportunity to study bacterial communities across different drinking water sources from around the world as these approaches become routine. Future studies should include the use of both culture-based and sequence-based approaches to better characterize the relationship between the two data types.

## Conclusions

Our controlled study provides compelling evidence of the efficacy of the PointONE™ filter in homes in a field setting, providing substantial reductions in bacterially-caused diarrhea for the entire length of our study (200 days post filter installation). Water quality sampling provided strong complementary evidence to support self-reported diarrhea information and supported the choice of this filtration technology that removes only microbial and particulate contaminants. While we observed no significant impact of school filters, in other contexts where water is being consumed more regularly by school children, additional impact may be observed. Further work is needed to evaluate the post-200 day utilization of filters and long-term filter efficacy in a field setting.

## Data Availability

The survey datasets used and/or analyzed during the current study are available from the corresponding author on reasonable request.
The 16S amplicon sequencing datasets generated and/or analyzed during the current study are available in the Short Read Archive (SRA), https://www.ncbi.nlm.nih.gov/bioproject/PRJNA670359. 

https://www.ncbi.nlm.nih.gov/bioproject/PRJNA670359

## Abbreviations

WHO: World Health Organization
EPA: Environmental Protection Agency

## Additional Files

Additional File 1 – AdditionalFile1.xlsx, Spreadsheet (.xlsx), List of water quality kits reported in this study, associated metadata for each kit, and water quality results.

Additional File 2 – AdditionalFile2.pdf, Document (.pdf), Supplemental Methods for water quality testing.

Additional File 3 – AdditionalFile3.pdf, Table (.pdf), Prevalence of self-reported diarrhea by home filter status: Intent to Treat Analysis.

Additional File 4 – AdditionalFile4.xlsx, Spreadsheet (.xlsx), Metadata file for 16S rRNA amplicon data analysis. The tab “DR_MiMarks-Metadata” contains all metadata associated with amplicon sequence data deposited in the SRA. The tab “Key” contains an explanation of all column headers.

## Declarations

### Ethics approval and consent to participate

Consent for participation in research was obtained from all participants. This study was conducted under the approval of the Dordt University Institutional Review Board.

### Consent for publication

Not applicable.

### Availability of data and materials

The survey datasets used and/or analyzed during the current study are available from the corresponding author on reasonable request.

The 16S amplicon sequencing datasets generated and/or analyzed during the current study are available in the Short Read Archive (SRA), https://www.ncbi.nlm.nih.gov/bioproject/PRJNA670359.

### Competing interests

Portions of the authors’ time (NT, KDVG, RDW, SAB, FSM, JWP, MJP and AAB) were supported by a grant from Sawyer Products, Inc.

### Funding

Portions of the authors’ time and materials were supported by a grant from Sawyer Products, Inc. Representatives of the funding agency were consulted during study design and aided in training for use of point-of-use filters in the field. Neither the funding agency nor its representatives contributed to data collection, data analysis or interpretation of data.

### Authors’ contributions

NT, KVDG, RU, RDW, SAB, AH and AAB co-led the development of the research questions and study design. DM, EB, OC, CGZ and NT led data monitoring, data cleaning and statistical analyses. AAB, SAB, MJP, JWP, BPK, ADS, RDW, FSM, TMB, CEAC, LE, BF, GKG, MAH, JAL, SML, TRL, JMP, ES, DJS, JES, MJS, MS, DRW designed laboratory protocols and water sampling kits, participated in RNA extraction, metals and particulate testing, final dataset creation and reporting. NT, KVDG, RU, RDW, SAB, BPK, and AAB wrote and revised the manuscript.

## Acknowledgments

The authors thank Darrel Larson, Chad Nelson, Gary Higgins and numerous others in the Child Hope Network for their efforts in administering data collection and their vision for the project.

## Notes

### Clinical Trial

NCT03972618

