## Additional File 2 for "Diarrhea prevalence in a randomized, controlled prospective trial of point-of-use water filters in homes and schools in the Dominican Republic"

### Additional File 2: Supplemental Methods

#### Sample Collection

##### *In Field Sample Collection*

Sampling to collect cells and particulate matter in source water was conducted with a tandem pair of 12 cm x 3.5 cm-diameter point-of-use 0.1 micron hollow fiber membrane filters (Sawyer Products, Inc.). The tandem unit was assembled by attaching two filters together with tubing; the first filter in the tandem pair was labeled “Filter A” and the second filter in the tandem pair was labeled “Filter B”. Field kits containing pre-labeled and pre-assembled tandem filter units, detailed instructions, and accessories to perform systematic sampling were assembled at Hope College, and delivered to the field collection sites in the Dominican Republic (Supplemental Figure 1). Clean 18.9 L (5 gallon) plastic buckets were fitted with filters and tubing. Buckets were rinsed with source water then filled with 16 L of source water which was allowed to gravity drain through the filter. After a bucket was drained, the tandem filter unit was detached and 4, 50 mL volumes of air were pushed through the sample with a syringe to flush out residual water. Each tandem filter unit was then capped at both ends, placed in a zip-sealed plastic bag and shipped back to Hope College. Each field kit contained materials for the collection of three biological replicates at each source water location. An image of a tandem filter setup in the field is shown in Supplemental Figure 1.

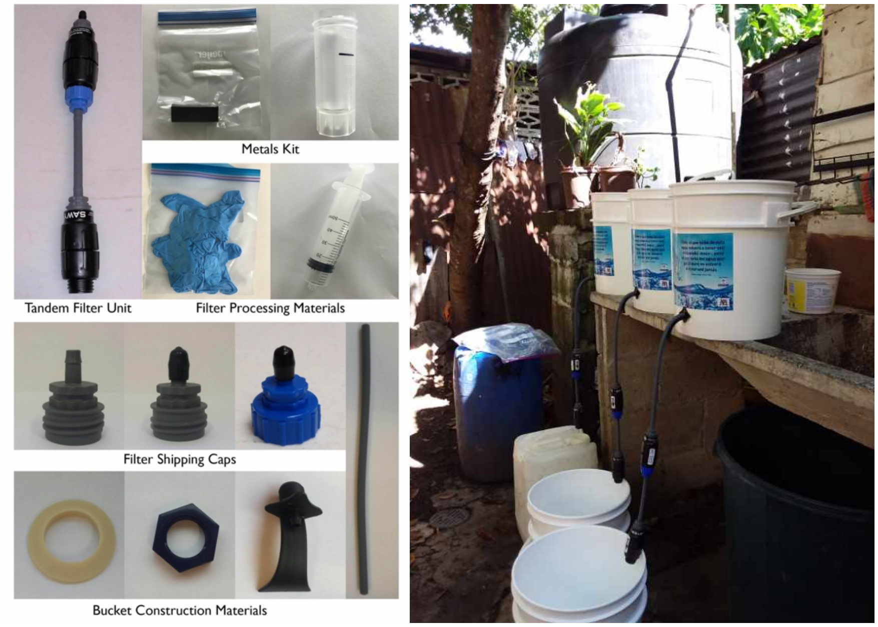


Figure S1. Kit components and field setup of tandem filter sampling of drinking water sources.

#### Sample Processing

##### *Collection of Particulates and Bacteria from Filters*

An apparatus (Supplemental Figure 2) was designed and constructed to backflush filters returned from the field in order to collect the source water particulates and bacterial cells captured on the filter. The apparatus was sterilized using a 70% ethanol solution each day prior to use and at the filter attachment point prior to attaching each filter. The air intake was fitted with a HEPA filter to prevent airborne contamination.

A filter was attached to the apparatus in a reverse orientation to filtering in the field. A slug of autoclaved 18MΩ water was flushed through the filter with 103 kPa air, and the sample containing removed particulates and bacteria was collected into a sterile 250 mL centrifuge bottle. Numerous trials with known amounts of material loaded onto filters determined that using two sequential slugs of 125 mL of water, at 103kPa air pressure, provided the highest systematic yield of filtered material (95 ± 5%).

Backflushed samples were centrifuged at 10,000 x G for 20 minutes to pellet cells and particulates. The supernatant was removed via vacuum aspiration and the pellet was resuspended in 30 mL of 18 MΩ water. A 200 µL aliquot of the homogenized pellet resuspension was pipetted directly into Qiagen DNeasy PowerViral lysis tubes and stored at -80C until DNA isolation was performed. The remaining pellet resuspension was used for particulate analysis.

For each day that backflushing was performed, controls were produced to assess the bacterial community associated with 1) the water used for backflushing (MQ Control, 200 µl directly into a DNA extraction tube; Blank Pellet, 250 mL centrifuged as above) and 2) unused filter devices (Control Pellet; 250 mL backflushed through a brand new filter).

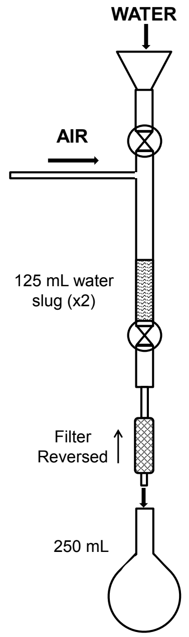


Figure S2. Schematic illustrating the laboratory back flushing apparatus to retrieve the suspended load particulates and cells captured by the filter at the drinking water source.

##### *Metal Retrieval*

Returned field foams were processed through an acid-wash procedure to recover adsorbed metals. Foam blocks were first dried at 100 ºC for 24 hours then weighed. Blocks were cut into two pieces and placed in a desiccator. Prior to rinsing, one half-block piece was reweighed and placed in an acid-washed 50 mL syringe equipped with a 0.45 µm syringe tip filter. 30 mL of 3% trace-metal grade nitric acid (pH~0.7) was added to the open syringe holding the foam, and a plunger was inserted. After 5 minutes the plunger was depressed and the acid was filter-pressed through the foam block into a 50 mL metal-free conical tube. Rinsates were analyzed immediately or refrigerated until analysis. Because the foam blocks employed for sequestering metals contained some of the target analyte metals as part of the chelating formulation, rinsates from background control foams were analyzed as well as controls of field, storage, and processing equipment.

***Amplicon Sequencing of 16S rRNA V4 Region***

DNA was extracted from backflushed material derived from filters using DNeasy PowerViral kits (MoBio Laboratories, Inc., CA, a Qiagen company) per the manufacturer's protocol. DNA samples were prepared for Illumina sequencing by following the MiSeq Wet Lab SOP [1]. Libraries for 94 DNA samples and two controls (a negative control and a positive, mock community control (ZymoBIOMICS Microbial Community Standard, Zymo Research, Irvine, CA) were processed in a 96 well plate; libraries for four, 96 well plates were combined for a single sequencing run. High throughput library preparation was performed with an EpMotion 5075 automated liquid handling device (Eppendorf North America, Enfield, CT). Amplification of the V4 region of the bacterial 16S rRNA gene was performed using universal primers (515F, 5'-GTGCCAGCMGCCGCGGTAA-3' and 907R 5'-CCGTCAATTCMTTTRAGTTT-3'; modified to include unique index pairs and adapters for Illumina sequencing) and Accuprime Pfx Supermix under the following conditions: 95 ˚C for 2 min; 30 cycles of 9 ˚C for 20 s, 55 ˚C for 15 s and 72 ˚C for 5 min; final elongation at 72 ˚C for 10 min. Amplicon size was checked on a 1% agarose gel and confirmed with High Sensitivity D1000 ScreenTape on the Agilent 2200 Tapestation system (Agilent Technologies Inc., CA). Amplicons were then purified using calibrated Ampure XP beads (Beckman Coulter; Brea, CA) and normalized with a SequelPrep kit (Invitrogen Corporation, CA). Normalized samples from a single 96 well plate were pooled into a single 1.5 mL tube. The pooled DNA was quantified using a Qubit dsDNA high sensitivity assay kit (Thermo Fisher Scientific Inc.; Waltham, MA). Four, pooled PCR amplicon libraries (equivalent of four, 96 well plates) were combined in equimolar concentrations into a single library. This combined library was sequenced using an Illumina MiSeq (500 cycle, 2 x 250bp paired-end v2 chemistry, Illumina, Inc., San Diego, CA) according to the manufacturer instructions.

#### Analysis

##### *Particulates-(Suspended Load)*

The suspended load present in source waters was estimated by resuspending backflush samples and immediately analyzing using a Microlab® FX522 spectrophotometry system. Attenuation (transmittance and absorbance) and scattering were measured at multiple wavelengths (refer to [2] for more detail). Particulate concentrations were estimated by comparison to standard curves of known suspended load that were developed by using individual common rock-forming minerals. These standard curves were considered representative of major types of geologic terrains, based on the assumption that the suspended load in any location is systematically reflective of the eroding substrate [3–6]. Many of these minerals were identified in the particulate material recovered upon back-flushing. Mineralogy was determined and/or estimated by powder X-ray diffraction (PXRD) techniques (Rigaku® MiniFlex+) and SEM-EDS (Hitachi® TM-3000) analysis. SEM-EDS was also used to identify elemental content of particulates.

*Metals in Rinsates-(Dissolved Metals)*

Rinsates were analyzed by ICP-OES techniques with a PerkinElmer® Avio 200 instrument. Quality assurance (QA) and quality control (QC) checks were consistent with a modified EPA Method 200.7 protocol, as summarized in methods provided by Perkin Elmer [7]. Analytes in the study include As, Se, Zn, Pb, Cd, Ni, Fe, Mn, Cr, Mg, Cu, Ce, Sb, and Ba. Additional information on detection wavelengths, ICP parameters and QA/QC for this study can be accessed in Peterson et al, 2020. Raw data were processed through a statistical comparison routine and reverse protocol algorithm to estimate metal concentrations in field drinking water sources. Several working assumptions were made for results obtained from metal retrieval and analysis procedures and element retention and recovery testing [2].

Single-element foam retention/recovery testing was performed to evaluate the relative validity of the working assumptions employed to estimate the heavy metal content in field source waters. Results from retention tests indicated a trend from higher recovery at low input solution concentrations to lower recoveries at high input solution concentrations. They also showed different recoveries for different elements. For example, within the 20-200 ppb range of input concentrations, the average recovery from all foams for Cu was: Cu = 241% (±210%). Cu is a high concentration background element within the polyurethane foams. These results substantiate the understanding that, except for Cu, metal concentrations determined in this study should be considered minimum levels. However, there are some cautionary considerations regarding the foam retention data. These are: 1) The tests were for single-metal solutions, the behavior of which may not reflect the processes active when multiple elements are present in solution with subsequent competition for adsorption sites on the foam; and, 2)The aqueous matrix of the test solutions (RO water) is not representative of the complex multi-constituent matrix of natural field waters.

As another check on the working assumptions, field tests on local lake water (Holland, MI, USA) were performed in which direct analysis of water was compared to analysis of rinsates from lake samples subjected to the field foam collection and rinsing protocol. Rinsates of foam-sequestered lake samples were significantly distinguishable from average background foam concentrations for As, Cd, Ni, Cr, Mg, Cu, Ce, Ba, and Se. However, only the concentrations of Mg, Ce, and Cu were significantly distinguishable when comparing direct lake samples with lake water rinsates after average background foam concentrations were subtracted. Mg concentrations were the only values above the LOQ. This experiment indicated that foam retention from actual field samples could only be calculated from sources with high enough concentration levels to distinguish from background, and further supports that field concentrations of most metals estimated in the current study should be considered minimum values.

*16S rRNA Communities*

Sequencing data from Illumina MiSeq runs were demultiplexed using the MiSeq Reporter Generate FASTQ workflow, producing forward and reverse read files in FASTQ format for each filter in a kit used to sample a drinking water source. Reads files from multiple sequencing runs were processed together according to the MiSeq SOP [8, 9] using mothur v.1.44.0 [10] via a batch script of mothur commands to produce a set of high quality operational taxonomic units (OTUs) at a similarity cutoff of 97%. Reads were aligned and taxonomy assigned using the Silva

Release 132 alignment and database [11]. Chimeras were removed from the OTUs using vsearch v2.13.3 [12]. After processing, the data set included 7,312,060 sequences, representing 88,319 unique sequences (OTUs).

The data set was analyzed using the *R* packages, Phyloseq [13] and vegan[14]. A phyloseq data object was generated from the mothur "shared" and "taxonomy" files and a metadata file describing characteristics of each sample. Each sequencing reaction is represented by a row in the metadata spreadsheet, with each row representing backflushed material from a single filter. Beta diversity analyses were performed by calculating Bray-Curtis pairwise distances followed by NMDS ordination (*k* ranging from 2 to 15, 200 iterations). The quality of sequencing reactions from each filter in each kit was initially evaluated by looking at the degree of clustering of filters from a kit in NMDS ordinations alongside control sequencing reactions associated with the date a kit was processed (backflush controls) and the library preparation plate the filters were processed on (plate controls). It was expected that the three Filter A replicates from a kit (water source) should cluster in the ordination and be separated from Filter B replicates of the same (and other) kit(s). The number of reads from a sequencing reaction were also used to evaluate the quality of the sequencing reactions. All sequencing reactions with fewer than 5000 reads were eliminated from further analysis. Five kits were eliminated from further analysis of bacterial communities (kits 221, 234, 241, 245, 247) after quality control evaluations were completed. PERMANOVA [15] and betadisper were performed on pairwise sample groupings based on these metadata categories: tandem_filter, filter, kit, water_type, and all subcategories of water_type.

In order to determine the presence/absence of 18 bacterial genera in backflush material from each filter, relative abundances of each OTU in a sample were calculated and OTUs were filtered to remove those below 0.1%. Each genus was considered present in a drinking water sample if the following criteria were met: 1) present in at least one Filter A replicate, 2) absent in all associated Filter B replicates, and 3) absent in all associated backflush controls and the negative plate control. Results for each water source (kit) are shown in Additional File 1.
