## Additional File 3 for "Diarrhea prevalence in a randomized, controlled prospective trial of point-of-use water filters in homes and schools in the Dominican Republic"

**Additional File 3. Prevalence of self-reported diarrhea by home filter status: Intent to Treat Analysis**

| Aggregation | Before filter in home[1] % (x/n) | After filter in home[1] % (x/n) | OR (95% CI)[2] | aOR (95% CI)[3] |
|---|---|---|---|---|
| Any member of the household | 25.6% (101/394) | 10.3% (70/681) | 0.26 (0.17,0.383)*** | 0.31 (0.18,0.52)*** |
| Adults only | 17.3% (68/392) | 4.8% (32/666) | 0.07 (0.03,0.16)*** | 0.26 (0.13,0.53)*** |
| School aged children only | 12.2% (46/378) | 5.7% (37/648) | 0.22 (0.09,0.5)*** | 0.41 (0.19,0.86)* |
| Children less than 5 | 11.8% (12/102) | 7.1% (12/169) | 0.50 (0.18,1.37) | 0.47 (0.12,1.83) |

*$p<0.05$; **$p<0.01$, ***$p<0.001$*

[1]Counts are each measurement of each household and include households reporting no filter usage

[2]Odds Ratio accounting for repeated measures

[3]Adjusting Odds Ratio accounting for repeated measures and adjusted for Household size, Season (Fall, Winter, Spring), Water Source, School Filter Status and Region
